# Safety and immunogenicity of a recombinant DNA COVID-19 vaccine containing the coding regions of the spike and nucleocapsid proteins: Preliminary results from an open-label, phase 1 trial in healthy adults aged 19–55 years

**DOI:** 10.1101/2021.05.26.21257700

**Authors:** Jin Young Ahn, Jeongsoo Lee, You Suk Suh, Young Goo Song, Yoon-Jeong Choi, Kyoung Hwa Lee, Sang Hwan Seo, Manki Song, Jong-Won Oh, Minwoo Kim, Han-Yeong Seo, Jeong-Eun Kwak, Jin Won Youn, Jung Won Woo, Eui-Cheol Shin, Su-Hyung Park, Young Chul Sung, Jun Yong Choi

## Abstract

**Background:** We investigated the safety and immunogenicity of two recombinant COVID-19 DNA vaccine candidates in first-in-human trials. GX-19 contains plasmid DNA encoding SARS-CoV-2 spike protein, and GX-19N contains plasmid DNA encoding SARS-CoV-2 receptor binding domain (RBD) foldon and nucleocapsid protein (NP) as well as plasmid DNA encoding SARS-CoV-2 spike protein.

**Methods:** Two open-label phase 1 trials of GX-19 and GX-19N safety and immunogenicity were performed in healthy adults aged 19–55 years. GX-19 trial participants received two vaccine injections (1·5 mg or 3·0 mg, 1:1 ratio) four weeks apart. GX-19N trial participants received two 3·0 mg vaccine injections four weeks apart.

**Findings:** Between June 17 and July 30 and December 28 and 31, 2020, 40 and 21 participants were enrolled in the GX-19 and GX-19N trials, respectively. Thirty-two participants (52·5%) reported 80 treatment-emergent adverse events (AE) after vaccination. All solicited AEs were mild except one case of moderate fatigue reported in the 1·5 mg GX-19 group. Binding antibody responses increased after vaccination in all groups. The geometric mean titers (GMTs) of spike-binding antibodies on day 57 were 85·74, 144·20, and 201·59 in the 1·5 mg, 3·0 mg GX-19 groups and the 3·0 mg GX-19N group, respectively. In GX-19N group, neutralizing antibody response (50% neutralizing titer using FRNT) significantly increased after vaccination, but GMT of neutralizing antibody on day 57 (37.26) was lower than those from human convalescent serum (288.78). GX-19N induced stronger T cell responses than GX-19. The magnitude of GX-19N-induced T cell responses was comparable to those observed in the convalescent PBMCs. GX-19N induced both SARS-CoV-2 spike- and NP-specific T cell responses, and the amino acid sequences of 15-mer peptides containing NP-specific T cell epitopes identified in GX-19N-vaccinated participants were identical with those of diverse SARS-CoV-2 variants

**Interpretation:** GX-19N is safe, tolerated and induces humoral and broad SARS-CoV-2-specific T cell response which may enable cross-reactivity to emerging SARS-CoV-2 variants.

**Funding:** This research was supported by Korea Drug Development Fund funded by Ministry of Science and ICT, Ministry of Trade, Industry, and Energy, and Ministry of Health and Welfare (HQ20C0016, Republic of Korea).

**Research in context:** *Evidence before this study:* To overcome the COVID-19 outbreak, the development of safe and effective vaccines is crucial. Despite the successful clinical efficacy of the approved vaccines, concerns exist regarding emerging new SARS-CoV-2 variants that have mutated receptor binding domains in the spike protein. We searched PubMed for research articles published up to May 1, 2021, using various combinations of the terms “COVID-19” or “SARS-CoV-2”, “vaccine”, and “clinical trial”. No language or data restrictions were applied. We also searched the ClinicalTrials.gov registry and World Health Organization (WHO) draft landscape of COVID-19 candidate vaccines for ongoing trials of COVID-19 vaccines up to May 1, 2021. Ten DNA-based vaccines, including the vaccine candidate reported here, are in ongoing clinical trials. Among these, safety and immunogenicity results were reported from only one phase 1 trial of a DNA vaccine against SARS-CoV-2 (INO-4800). INO-4800 demonstrated favorable safety and tolerability and was immunogenic, eliciting humoral and/or cellular immune responses in all vaccinated subjects. There is only one ongoing clinical trial of a vaccine against SARS-CoV-2 variants (mRNA-1273.351).

*Added value of this study:* This is the first-in-human phase 1 trial in healthy adults of a recombinant DNA vaccine for COVID-19 (GX-19N) containing the coding regions of both the spike and nucleocapsid proteins. This trial showed that GX-19N is safe, tolerated, and able to induce both humoral and cellular responses. A two-dose vaccination of 3·0 mg GX-19N (on days 1 and 29) induced significant humoral and cellular responses. The neutralizing geometric mean titers in individuals vaccinated with GX-19N were lower than those of human convalescent sera. However, the GX-19N group showed increased T cell responses, which was similar to those analyzed using convalescent PBMCs. Furthermore, GX-19N induced not only SARS-CoV-2 spike-specific T cell responses but also broad nucleocapsid-specific T cell responses, which were also specific to SARS-CoV-2 variants.

*Implications of all the available evidence:* It is important to note that GX-19N contains a plasmid encoding both the spike and nucleocapsid proteins, and that it showed broad SARS-CoV-2-specific T cell responses, which may allow cross-reactivity with emerging SARS-CoV-2 variants. Based on these safety and immunogenicity findings, GX-19N was selected for phase 2 immunogenicity trials.

## Introduction

To overcome COVID-19 outbreak, development of safe and effective vaccines is crucial. Many are in development based on different platforms including mRNA, DNA, viral vectored, protein-based and inactivated virus vaccines. Most vaccines that are currently in development target the spike protein, which binds to the angiotensin converting enzyme 2 receptor to entry into the host cell.

Despite the successful clinical efficacy of the approved vaccines, the emergence of new variants possessing mutations in the receptor binding domain (RBD) of the S protein result in reduced neutralization by therapeutic antibodies and vaccine-induced immune sera^1,2^, in line with the reduced clinical efficacies of the approved vaccines in South Africa.^3^ While the development of new vaccines against these variants has already started (Moderna, NCT04785144), SARS-CoV-2 variants are likely to rapidly evolve into escape variants through mutations in the RBD, as illustrated in the evolution of human coronavirus 229E over time.^4^ Virus-specific T cells can be an alternative to neutralizing antibodies against variants. Given that variants escaping T cell immunity against SARS-CoV-2 also occur^5^, broader T cell immunity would be beneficial and can be achieved by adding well-conserved antigens to induce dominant T cell responses.

GX-19 is a candidate recombinant DNA vaccine that contains a plasmid encoding the SARS-CoV-2 S proteins (S1, S2).^6^ A plasmid vector (pGX27), which was used for the HPV DNA vaccine undergoing clinical trials, was used.^7,8^ We produced the following four prototype DNA vaccines expressing the SARS-CoV-2 S protein: 1) S1S2 (GX-19A), 2) S1 (GX-19B), 3) S1S2 with the CD40 ligand (GX-19C), and 4) S1 with the CD40 ligand (GX-19D). Preclinical studies in mouse and monkey models showed that these four vaccine candidates can induce neutralizing antibodies and cytotoxic T cell responses. Among them, GX-19A was selected (GX-19) because it induced the highest titer of SARS-CoV-2-specific neutralizing antibodies and cytotoxic T cell responses.

Another candidate vaccine is GX-19N, a recombinant DNA vaccine that encodes the S protein, RBD-foldon, and nucleocapsid protein (NP). GX-19N is composed of the GX-19 plasmid (pGX27-S1S2) and a plasmid vector expressing the RBD-foldon with NP (pGX27-SRBD/NP) in a ratio of 1:2 (Figure 1). Preclinical studies in mouse and monkey models showed that higher neutralizing antibody responses and cellular immune responses were expected following vaccination with GX-19N. Details regarding the results of preclinical studies are provided in the Supplementary Appendix 1. Here, we report the preliminary safety and immunogenicity assessments of two recombinant DNA vaccine candidates for COVID-19, GX-19 and GX-19N, in first-in-human trials.

**Figure 1.**
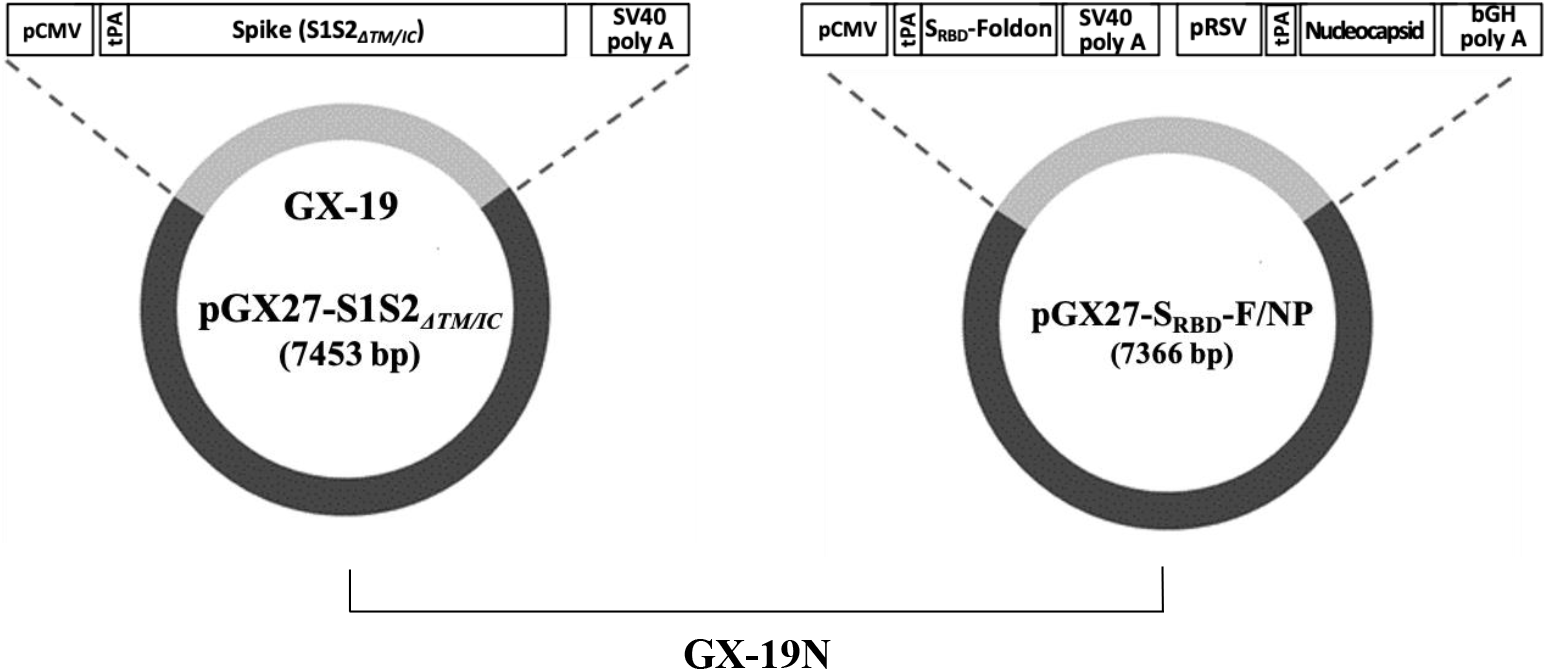
Diagram of the GX-19 and GX-19N DNA vaccines. GX-19 (pGX27-S1S2ΔTM/IC) contains the SARS-CoV-2 spike (S) gene lacking the transmembrane (TM)/intracellular (IC) domain, constructed by inserting the gene sequence into the pGX27 vector. GX-19N consists of GX-19 and a second plasmid, pGX27-SRBD-F/NP at a 1:2 ratio. pGX27-SRBD-F/NP is designed to express the fusion protein of the receptor binding domain (RBD) of the spike protein and the T4 fibritin C-terminal foldon (SRBD-foldon) and nucleocapsid protein (NP). The S, SRBD-foldon, and NP sequences are preceded by the secretory signal sequence of the tissue plasminogen activator (tPA). pCMV, cytomegalovirus early enhancer/promoter; SV40 poly A, Simian virus 40 late polyadenylation sequence; pRSV, Rous sarcoma virus promoter; bGH polyA, bovine growth hormone polyadenylation sequence.

## Methods

### Study design and participants

We performed two open-label phase 1 trials to evaluate the safety, tolerability, and immunogenicity of GX-19 (NCT04445389) and GX-19N (NCT04715997) after intramuscular vaccination of healthy adults at two hospitals in South Korea (Severance Hospital and Gangnam Severance Hospital). The GX-19 trial was a dose-escalation study conducted prior to the GX-19N trial. GX-19 trial participants received two injections of the trial vaccine four weeks apart at a dose of 1·5 mg or 3·0 mg. The GX-19N trial was subsequently conducted based on the results obtained in the GX-19 trial.

Healthy adult participants aged 19–55 years were enrolled through local advertisements. Full details of the eligibility criteria are described in the trial protocol provided in the Supplementary Appendix 2.

Written informed consent was obtained from all participants. The trials were conducted in accordance with the principles of the Declaration of Helsinki and Good Clinical Practice and registered with ClinicalTrials.gov (NCT04445389 and NCT04715997). This study was approved in South Korea by the Korea Ministry of Food and Drug Safety (reference 20200261849) and the Institutional Review Board of Severance Hospital (IRB No. 4-2020-1220).

### Procedures

The vaccines were produced according to current good manufacturing practices and supplied by BINEX Co., Ltd., Korea, a contract manufacturing organization. The vaccines were delivered using an electroporator (Elimtek Co., Ltd., Korea) to maximize cellular DNA uptake by generating a high-voltage pulse after drug injection.

In the GX-19 trial, participants were first enrolled in the 1·5 mg group. After safety and tolerability data from this group were reviewed by the independent Data Safety Monitoring Committee (DSMC), participants were enrolled in the 3·0 mg group. Participants were administered 1·5 mg (0·38 ml) or 3·0 mg (0·75 ml) GX-19 intramuscularly in the deltoid muscle on days 1 and 29 and followed for 52 weeks post first vaccination. In the GX-19N trial, participants were enrolled in a single dose group to receive 3·0 mg (0·75 ml) GX-19N intramuscularly in the deltoid muscle on days 1 and 29 and followed for 52 weeks post first vaccination. To assure participant safety, all participants were followed-up with daily by telephone for AEs for up to seven days from the first vaccination and had to visit the site on the eighth day to undergo a safety assessment.

Follow-up visits were scheduled on days 8, 29, 43, 57, and weeks 24 and 52 after the first vaccination, and safety-related data were collected. A diary was distributed to participants at the time of the first and second vaccination for them to personally list any AEs experienced over approximately four weeks. To assess immunogenicity for this interim analysis report, blood samplings were performed at baseline, 29, 43, and 57 days after the first vaccination. Additional details regarding these procedures are provided in the Supplementary Appendix 3.

### Safety

Local and systemic reactions after each vaccination were monitored for 28 days. Serious AEs were recorded throughout the follow-up period. The solicited AEs appearing in the study participants were graded according to the “Guidance for Adverse Event Grading Scale for Volunteers in Preventive Vaccine Clinical Trials (2011, 12)” issued by the Ministry of Food and Drug Safety.^9^ The severity of unsolicited AEs was assessed according to Common Terminology Criteria for Adverse Events (CTCAE V5.0)^10^ and Medical Dictionary for Regulatory Activities (MedDRA) classification and relatedness to the vaccine. AEs of special interest (AESIs), including acute respiratory distress syndrome, pneumonitis, enhanced disease following immunization, acute cardiac injury, arrhythmia, septic shock-like syndrome, and acute kidney injury, were also assessed. Laboratory AEs were graded using site-specific toxicity tables, which were adapted from the guideline for assessing the severity of AEs in vaccine clinical trials by the Korea Ministry of Food and Drug Safety.^9^

### Immunogenicity

Binding antibody responses against the SARS-CoV-2 S protein were assessed by using an in-house-developed ELISA and antibodies against the SARS-CoV-2 RBD protein were assessed using an ELISA kit (Bionote Inc, Korea). Binding antibodies were assessed before the vaccination, and at days 43 and 57 after the first vaccination. Vaccine-induced neutralizing activity was assessed using a live wild-type SARS-CoV-2 focus reduction neutralization testing (FRNT) assay and a SARS-CoV-2 Spike (S)-pseudotyped murine leukemia virus (MLV) retrovirus-based single-round-of-infection neutralization assay (PsVNA). The 50% neutralization titers were reported as the interpolated reciprocal of the dilutions yielding 50% reductions in viral foci. The FRNT assay was performed on specimens collected before the vaccination and on days 43 and 57 after the first vaccination. For this interim analysis report, PsVNA results were available only before and on day 43 after the first vaccination. T cell responses in participants vaccinated with GX-19 or GX-19N were evaluated before and on day 43 after the first vaccination using a direct *ex vivo* IFN-γ enzyme-linked immunosorbent spot (ELISpot) assay with peripheral blood mononuclear cells (PBMCs). In these assays, PBMCs were stimulated overnight with overlapping peptides (OLPs) encoding the full-length sequence of S (for GX-19) and S/NP (for GX-19N), respectively. To selectively evaluate vaccine-induced SARS-CoV-2-specific T cell responses, the T-SPOT assay (Oxford Immunotec, UK) was performed. For this assay, PBMCs obtained from participants were stimulated with OLPs spanning S and NP that specific sequences with high homology to endemic (non-SARS-CoV-2) coronaviruses removed. The details of all methods are described in the Supplementary Appendix 4.

To compare vaccine-induced immune responses with those induced by natural SARS-CoV-2 infection, we analyzed convalescent blood specimens collected within three months of polymerase chain reaction-based diagnosis of COVID-19. FRNT/PsVNA and ELISpot assays were performed to evaluate neutralizing antibody responses and T cell responses, respectively.

### Outcomes

The primary outcome was the safety and tolerability of GX-19 and GX-19N based on observation of the frequency, characteristics, and severity of AEs experienced within 28 days of each vaccination. The secondary outcome was the immunogenicity of GX-19 and GX-19N. Immunogenicity endpoints in this interim analysis included changes in SARS-CoV-2 S and RBD antigen-specific binding antibody levels compared to baseline levels within day 57 in all participants (on days 1, 43, and 57 after first vaccination) and changes in SARS-CoV-2 neutralizing antibody levels within day 57 in the participants of the 3·0 mg GX-19N group (on days 1, 43 and 57 for the FRNT assay and day 1 and 43 after first vaccination for PsVNA). Immunogenicity endpoints also included determination of the SARS-CoV-2-specific T cell responses of GX-19 and GX-19N measured on days 1 and 43 after the first vaccination.

### Statistical analysis

Continuous variables are presented using descriptive statistics (sample size, mean, standard deviation, median, and range), and categorical variables are presented as the frequency and percentage, as well as a 95% two-sided confidence interval (CI), as appropriate. When theoretical hypothesis testing was required, parametric methods were used for continuous values with normal distribution and non-parametric methods were used for continuous values with non-normal distribution. Chi-square and Fisher’s exact tests were used for categorical values.

For safety assessment, AEs are presented as the number and percentage of subjects who experience AEs and the number of actual cases with a 95% CI. AEs were classified by type, severity, and causality for each group, and the frequency and percentage of each was calculated.

For immunological analysis, the GMTs of binding and neutralizing antibodies before and after the vaccination were assessed, and a 95% CI was presented. The proportions of participants with a ≥4-fold increase in the titer of binding and neutralizing antibodies compared to baseline levels were also presented. T cell responses were expressed as the number of spot-forming-units per 10^6^ cells measured using IFN-γ ELIspot and T-SPOT assays before and after the vaccination, and a 95% CI was presented. The subjects with ≥2-fold or ≥100 spots/10^6^ PBMCs increase in IFN-γ ELIspot assay on day 43, compared to day 1, were regarded as responders. The analyses were performed using IBM SPSS statistics and GraphPad Prism, version 8.4.1., and 95% CIs were calculated using the Clopper-Pearson method.

## Results

### Participant demographics

In the GX-19 trial, 96 healthy individuals were screened and 40 were enrolled between June 17 and July 30, 2020, to receive two doses of 1·5 mg or 3·0 mg GX-19 in an 1:1 ratio. In the GX-19N trial, 23 healthy individuals were screened and 21 were enrolled between December 28 and December 31, 2020 and received two doses of 3·0 mg GX-19N, (Figure 2).

**Figure 2.**
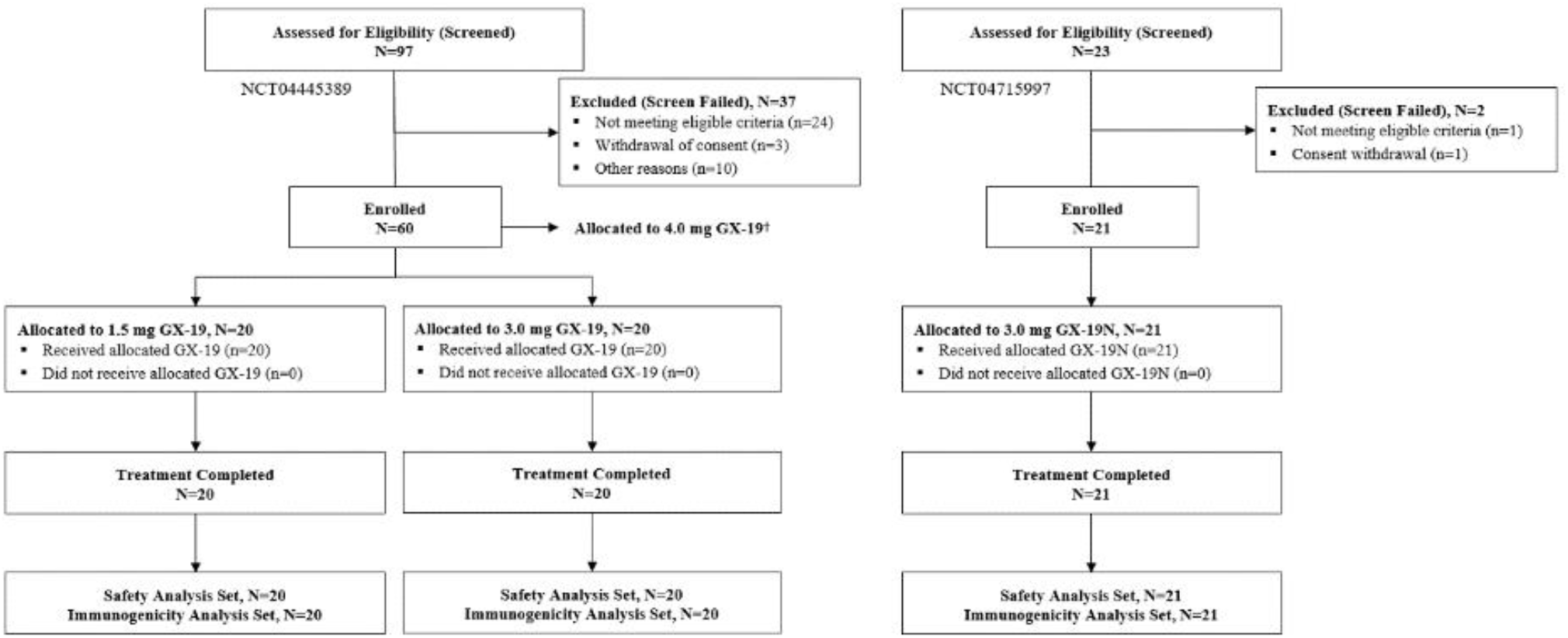
Trial flow diagram. †Twenty subjects were allocated to receive 4·0 mg GX-19 using a Jet Injector (data not shown).

All 61 participants received their second vaccine dose, and all continued to attend the scheduled trial visits until day 57 and were included in both safety and immunogenicity analyses. The demographic characteristics of the participants at enrollment are presented in Table 1. The mean age was 34·9 years (range, 20–52 years) and 52·4% were men. All participants were Korean. Similar demographic characteristics were observed in all groups.

**Table 1.**
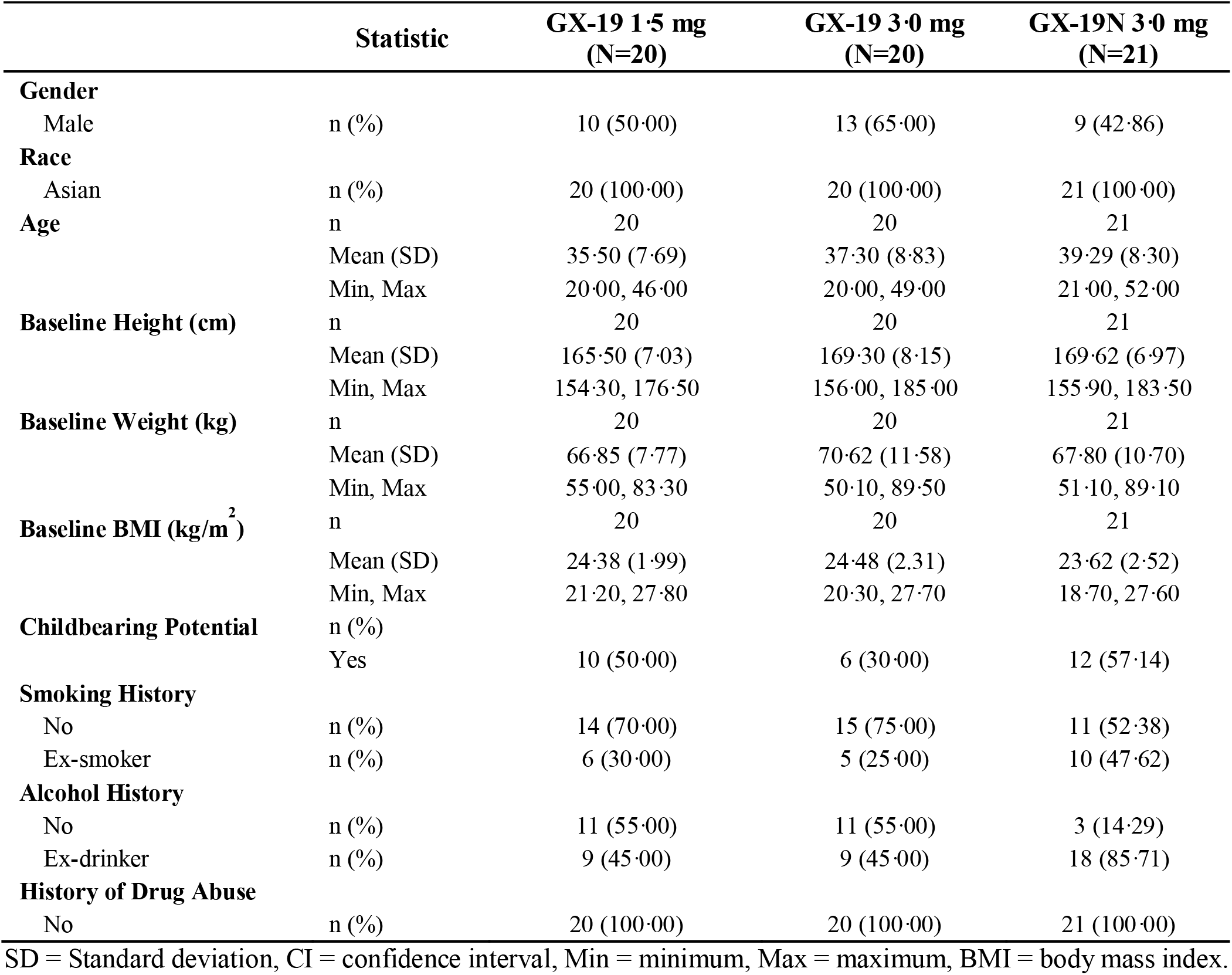
Descriptive statistics of demographic characteristics.

### Safety and tolerability

Thirty-two participants (52·5%), of whom 12 (60%) were in the 1·5 mg GX-19 group, eight (40%) in the 3·0 mg GX-19 group, and 12 (57·1%) in the 3·0 mg GX-19N group, reported 80 treatment-emergent adverse events (TEAEs) after vaccination. (Supplementary Appendix 5) The majority of AEs were mild, and none of the participants experienced serious drug-related AEs or AESIs. (Supplementary Appendix 6) No participants discontinued the trial due to AEs.

After the first vaccination, solicited local AEs were reported by six participants (30%) in the 1·5 mg GX-19 group, two (10%) in the 3·0 mg GX-19 group, and five (23·8%) in the 3·0 mg GX-19N group. After both doses, solicited local AEs were reported by six (30%) participants in the 1·5 mg GX-19 group, three (15%) in the 3·0 mg GX-19 group, and five (23·8%) in the 3·0 mg GX-19N group. Solicited systemic AEs were reported by one (5%) participant in the 1·5 mg GX-19 group and two (9·5%) in the 3·0 mg GX-19N group after the first vaccination and three (15%) in the 1·5 mg GX-19 group and two (9·5%) in the 3·0 mg GX-19N group after both doses. There were no reported solicited systemic AEs in the 3·0 mg GX-19 group. The most common solicited AEs were injection site pain, itching, tenderness, and fatigue. Solicited local and systemic AEs are shown in Figure 3. All solicited AEs developed within two days from vaccination and most events resolved within three days of onset. All AEs were mild except one case of moderate fatigue reported in the 1·5 mg GX-19 group. There was no increase in the number of solicited AEs related to the vaccine dose. There was no increase in the frequencies of AEs with the second vaccination compared to those of the first vaccination in all groups.

**Figure 3.**
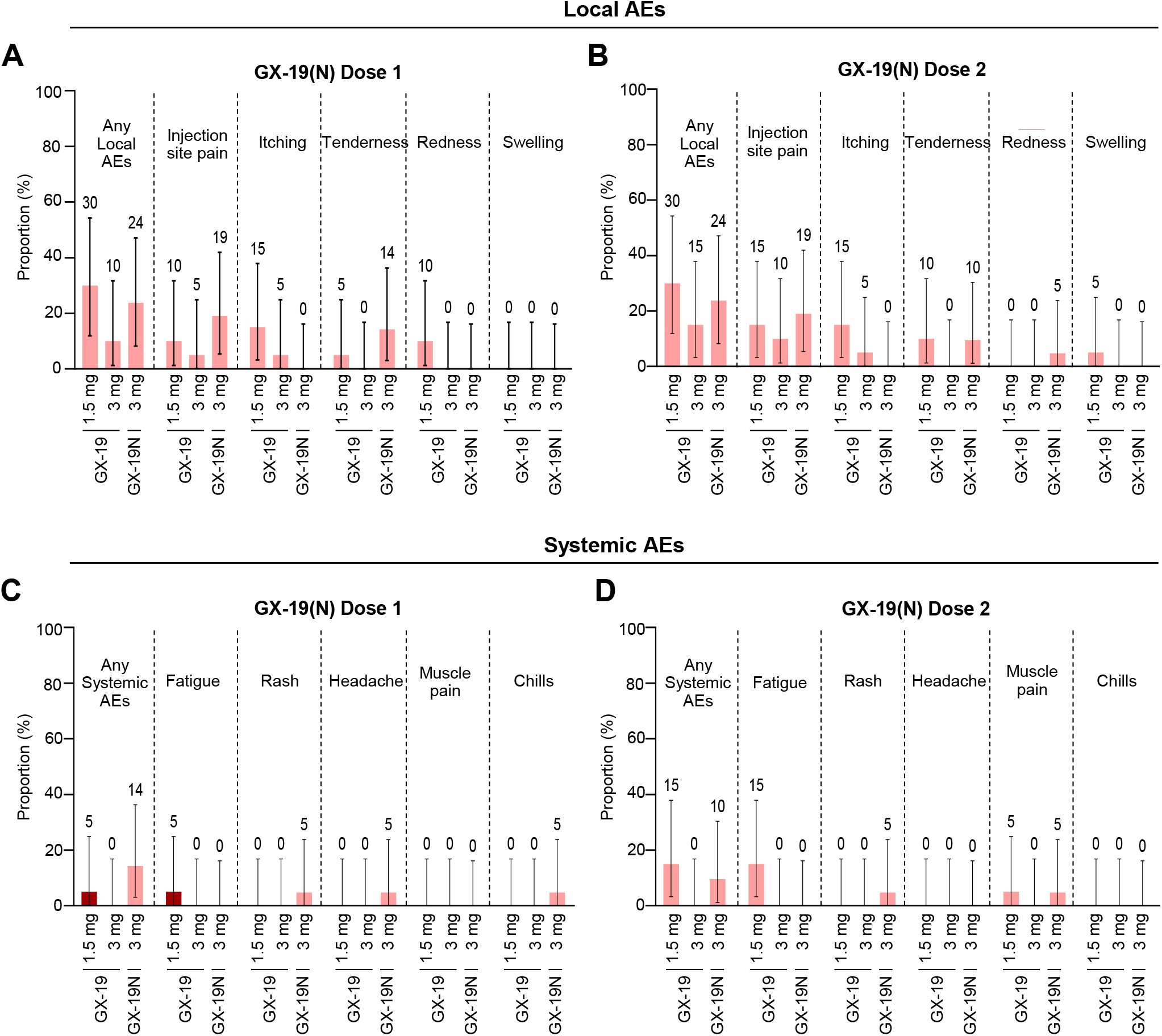
Solicited adverse events (AEs) reported after the administration of GX-19(N). The percentage of participants with AEs during the 28-day post vaccination period in each vaccine group (GX-19 1·5 mg, GX-19 3·0 mg, and GX-19N 3·0 mg) is plotted for solicited local (A, B) and systemic (C, D) AEs. There were no severe events. Note: Error bars represent the 95% confidence intervals.

Twenty-eight unsolicited AEs were reported by 16 participants. Most were predominantly mild and distributed similarly across the groups (four in the 1·5 mg GX-19 group, six in the 3·0 mg GX-19 group, and six in the 3·0 mg GX-19N group). There were two severe unsolicited AEs in two participants, namely, acute appendicitis in the 3·0 mg GX-19 group and acute cholecystitis in the 3·0 mg GX-19N group. Both events were unlikely to be related to the vaccine. Vaccine-related AE (transient mild paresthesia) was reported by only one participant in the 1·5 mg GX-19 group. Changes in laboratory variables (mild and transient elevation in the levels of aspartate aminotransferase and alanine aminotransferase) were observed in one participant in the 3·0 mg GX-19N group.

### Immunogenicity

#### SARS-CoV-2 binding antibody responses

The GMTs of binding antibodies to S protein increased after the first vaccination by day 57 in both the 1·5 mg GX-19 and 3·0 mg GX-19 groups. In the GX-19N group, antibody responses to S and RBD proteins increased after vaccination by day 43, but the GMTs on day 57 slightly decreased compared to those on day 43 (Table 2, Figure 4A). In each group, there was a statistically significant increase in GMTs by day 43 and day 57 over the baseline. The GMTs of S-binding antibody on day 57 were 85·74 (95% CI 49·67–148·00) in the 1·5 mg GX-19 group, 144·20 (95% CI 80·81–257·32) in the 3·0 mg GX-19 group, and 201·59 (105·32–385·83) in the 3·0 mg GX-19N group. The proportion of subjects with ≥4-fold rises in the S or RBD protein antibody levels after the vaccination was 55% in the 1·5 mg GX-19 group, 65% in the 3·0 mg GX-19 group, and 80·95% in the 3·0 mg GX-19N group.

**Table 2.** Summary of the spike and receptor-binding domain (RBD) binding antibody titers in serum after dosing with GX-19(N)

| Statistic |  | GX-19 1·5 mg<br>(N=20) | GX-19 3·0 mg<br>(N=20) | GX-19N 3·0 mg<br>(N=21) |
| --- | --- | --- | --- | --- |
|  |  | Spike | Spike | Spike/RBD |
| **Day 1** | N | 20 | 20 | 21 |
|  | GMT (SD) | 25·49 (2·6) | 33·64 (3·00) | 24·38 (3·67) / 1·14 (1·83) |
|  | 95% CI | 16·25, 39·98 | 20·11, 56·25 | 13·49, 44·05 / 0·87, 1·50 |
|  | Mean | 42·50 | 61·00 | 97·14 / 1·71 |
|  | Median | 20·00 | 30·00 | 20·00 / 1·00 |
|  | Min-Max | 10·00-160·00 | 10·00-320·00 | 10·00-1280·00 / 1·00-16·00 |
| **Day 43** | n | 20 | 20 | 21 |
|  | GMT (SD) | 60·63 (3·11) | 129·96 (3·58) | 341·84 (4·78) / 8·55 (7·18) |
|  | 95% CI | 35·67, 103·05 | 71·59, 235·93 | 167·75, 696·60 / 3·48, 20·97 |
|  | ≥4-fold increase, (%) | 35·00 | 65·00 | 80·95 |
|  | Mean | 107·50 | 255·00 | 726·19 / 114·90 |
|  | Median | 80·00 | 160·00 | 640·00 / 8·00 |
|  | Min-Max | 10·00-640·00 | 20·00-1280·00 | 10·00-2560·00 / 1·00-2048·00 |
|  | p-value [1] | 0·0010 | <0·0001 | <0·0001 / <0·0001 |
| **Day 57** | n | 20 | 20 | 21 |
|  | GMT (SD) | 85·74 (3·21) | 144·20 (3·45) | 201·59 (4·16) / 5·38 (6·31) |
|  | 95% CI | 49·67, 148·00 | 80·81, 257·32 | 105·32, 385·83 / 2·33, 12·45 |
|  | ≥4-fold increase, (%) | 55·00 | 65·00 | 80·95 |
|  | Mean | 166·00 | 286·00 | 433·33 / 62·00 |
|  | Median | 120·00 | 160·00 | 160·00 / 4·00 |
|  | Min-Max | 10·00-1280·00 | 20·00-1280·00 | 10·00-2560·00 / 1·00-1024·00 |
|  | p-value [1] | 0·0002 | <0·0001 | 0·0004 / 0·0001 |

GMTs = geometric mean titers, SD = Standard deviation, CI = confidence interval, Min = minimum, Max = maximum.
[1] p-value was calculated in Wilcoxon signed rank test.
Denominator of percentage is the number of subjects in each cohort.

**Figure 4.**
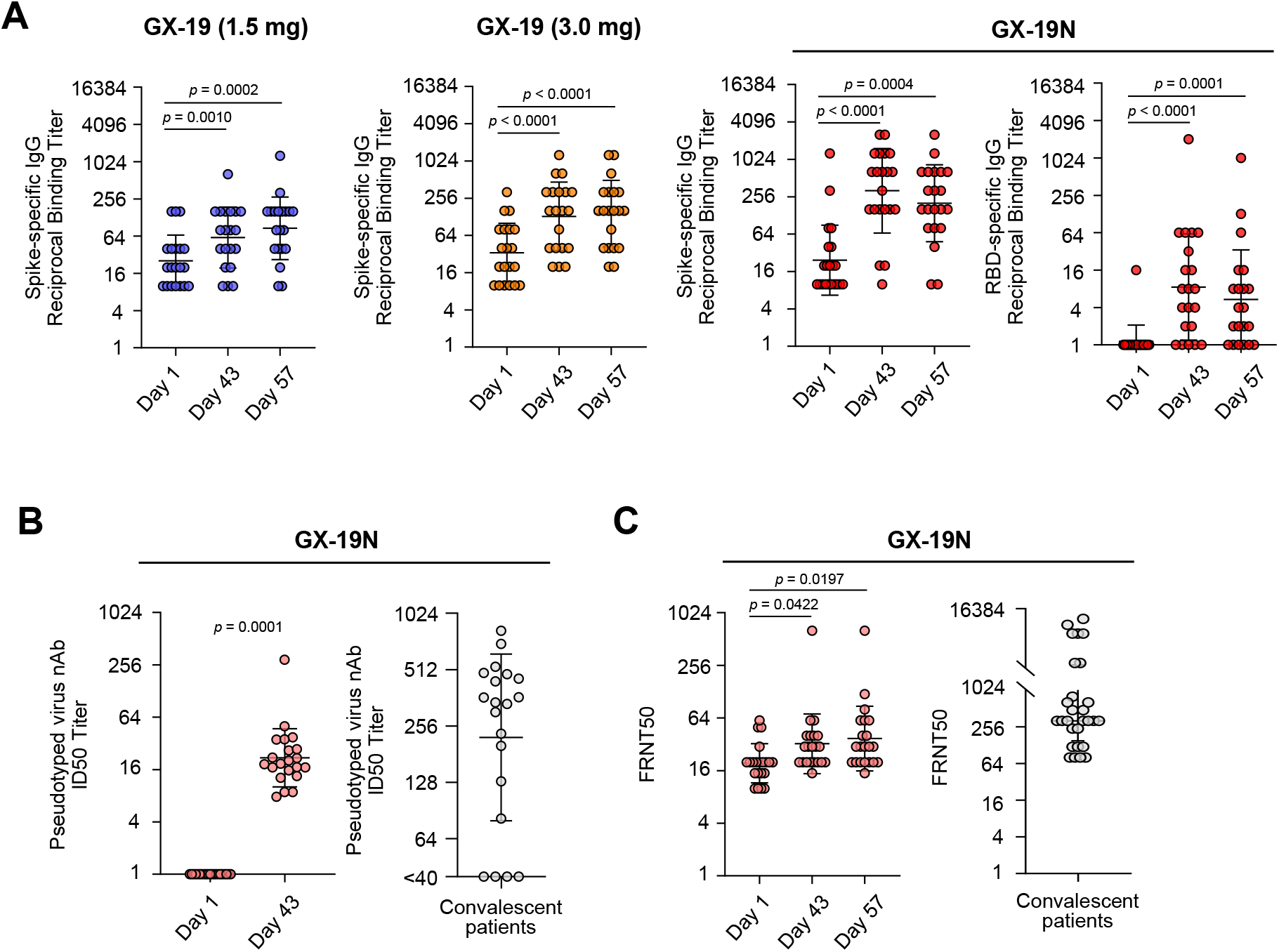
GX-19N DNA vaccine induced binding and neutralization antibody responses. Healthy volunteers aged 19-55 were vaccinated at day 1 and 29, with 1·5 mg (n=20), 3·0 mg (n=20) GX-19 or 3·0 mg GX-19N (n=21). Sera were collected at day 1, 43, 57 and assessed for SARS-CoV-2 Spike- and RBD-specific IgG antibodies by ELISA (A). GX-19N-induced neutralization antibody responses were determined by pseudotype virus neutralization assay (PsVNA) (B) and focus reduction neutralization test (FRNT) (C). Each circle indicates reciprocal binding antibody titer, ID50 (50% inhibitory dilution) or FRNT50 titer of each serum sample. IC50 and FRNT50 titers of serum samples obtained from convalescent patients are shown in the lower right panel. *p* values were calculated by using the Wilcoxon matched-pairs signed rank test.

#### SARS-CoV-2 neutralization responses

Based on the results of binding antibody responses, neutralization responses were only analyzed in the 3·0 mg GX-19N group. To demonstrate the neutralizing activity of GX-19N-induced antibodies, *in vitro* neutralization was assessed using FRNT with a live wild-type SARS-CoV-2 and PsVNA (Figure 4B and 4C). No participants had detectable PsVNA responses before the vaccination and a statistically significant increase in PsVNA responses was observed after the vaccination. The GMTs of the 50% inhibitory dilution (ID50) values in PsVNA were 1·00 (95% CI 1·00–1·00) before the first vaccination and 21·87 (95% CI 15·30–31·07) on day 43. Higher than 4-fold rises in the 50% neutralization titer were observed in all participants using PsVNA. However, the GMT of ID50 values on day 43 was lower than that of convalescent sera from 20 patients (GMT = 222·20) (Figure 4B). Using FRNT, a statistically significant increase in the neutralizing antibody GMT after vaccination was observed in the 3·0 mg GX-19N group by days 43 and 57. Neutralizing antibody GMTs were 32·47 (95% CI 22·71–46·41) on day 43 and 37·26 (95% CI 25·29–54·90) on day 57. The proportion of subjects with ≥4-fold rises in antibody levels was 23·81% by day 57. Neutralizing GMTs by day 57 were also much lower than those in 22 human convalescent serum samples (GMT = 288·78) (Figure 4C).

#### SARS-CoV-2-specific T cell responses

In the GX-19 trial, both the 1·5 mg and 3·0 mg GX-19 groups showed significantly enhanced S-specific T cell responses following vaccination. Specifically, 10 (50%) participants in the 1·5 mg GX-19 group and 10 (50%) in the 3·0 mg GX-19 group mounted augmented S-specific T cell responses on day 43, respectively (Table 3 and Figure 5A). In the GX-19N trial, GX-19N-immunized participants exhibited significantly enhanced S- and NP-specific T cell responses on day 43 after vaccination (Table 3, Figure 5B and 5C). The results revealed that 15 (75%) participants generated vaccine-induced T cell responses after vaccination (Supplementary Appendix 7), and the magnitude of GX-19N-induced T cell responses was comparable to those observed in the convalescent PBMCs from six COVID-19 patients (Figure 5B). Furthermore, we tried to evaluate whether five participants (19Na, 19Nd, 19Nf, 19Nq, and 19Nt), who did not show enhanced T cell responses based on the results of the IFN-γ ELIspot assay using OLPs spanning the full length of S and NP, induced vaccine-induced SARS-CoV-2-specific T cell responses. To this end, we used the T-SPOT kit with PBMCs from four participants (19Na, 19Nd, 19Nf, and 19Nq) using OLPs that did not contain specific sequences with high homology to endemic (non-SARS-CoV-2) coronaviruses. The results revealed that three out of four participants mounted increased GX-19N-induced SARS-CoV-2 S- and/or NP-specific T cell responses (Supplementary Appendix 8), indicating that a significant portion of participants classified as non-responders actually generated vaccine-induced T cell responses.

**Table 3.**
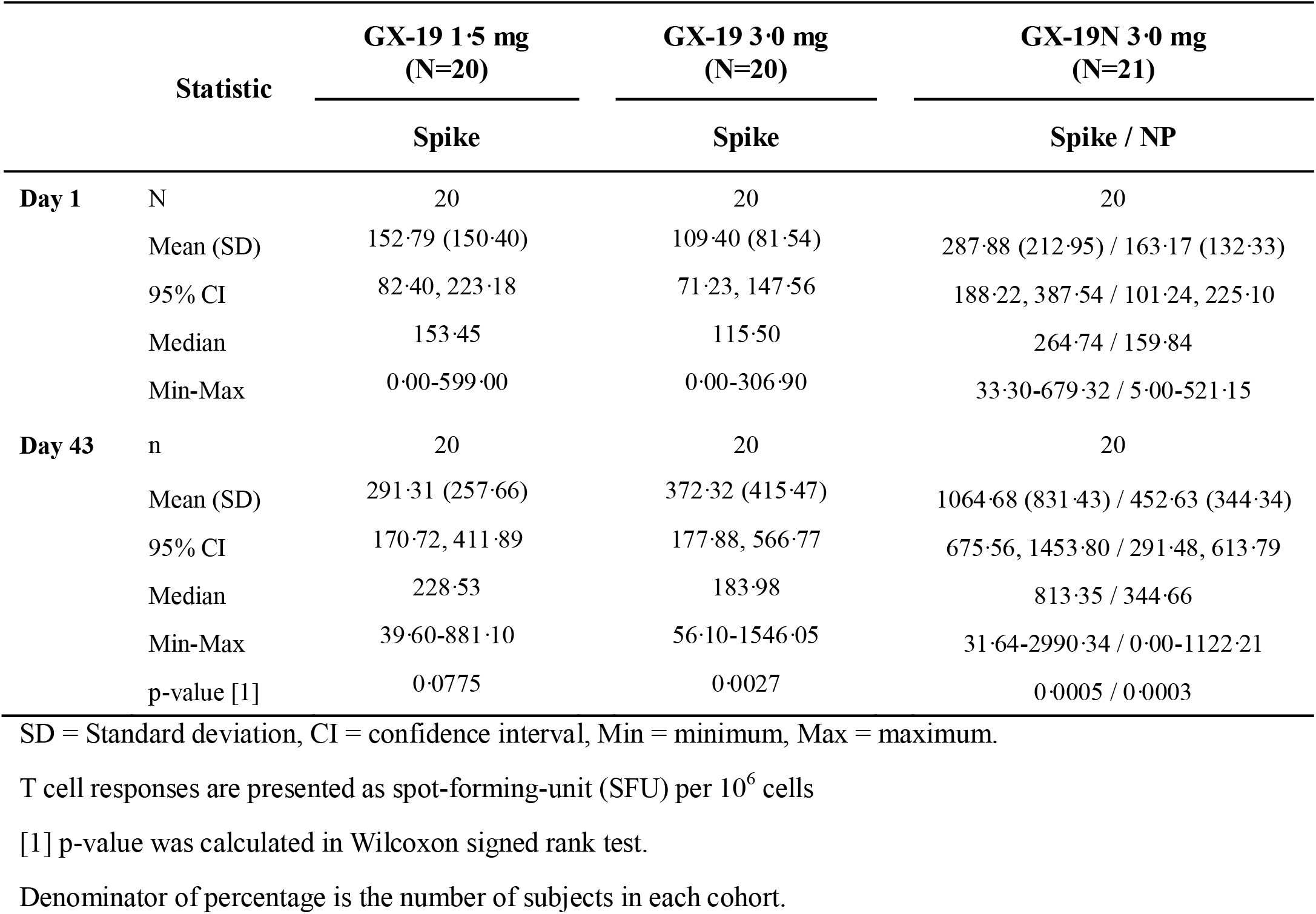
Summary of the specific T-cell response in serum after dosing with GX-19(N)

**Figure 5.**
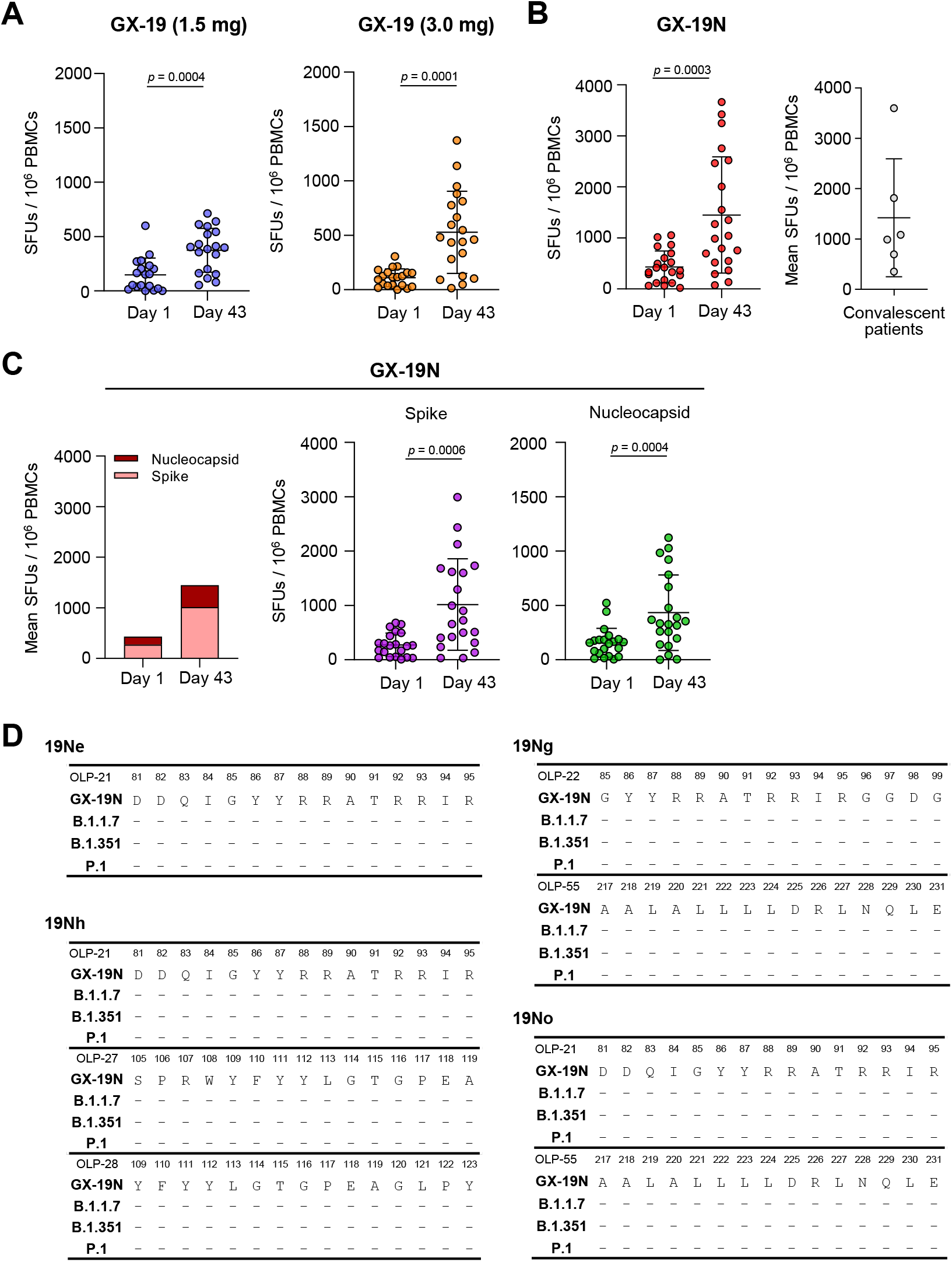
GX-19N elicited robust T-cell responses to SARS-CoV-2 antigens. Antigen-specific T cell responses at day 1 and day 43 were measured by IFN-γ ELISPOT assay using peripheral blood mononuclear cells (PBMCs) stimulated with overlapping peptides (OLPs) spanning full length of spike (S) (for GX-19) and S and nucleocapsid protein (NP) (for GX-19N). Individual data points are shown as a scatter dot plot with lines showing the mean with standard deviation. S-specific T cell responses at day 1 and day 43 in GX-19-immunized participants (A). Sum of S and NP-specific (B), S-specific or NP-specific (C) T cell responses at day 1 and day 43 in GX-19N-immunized participants are represented. Comparison of amino acids sequence of 15-mer peptides containing NP-specific epitopes for GX-19N-vaccinated participants with those of SARS-CoV-2 variants such as B.1.1.7, B.1.351 and P1(D). SFU, spot-forming-unit.

We next examined whether the NP-specific T cell responses induced by GX-19N were also specific to the NP of SARS-CoV-2 variants, such as B.1.1.7 (United Kingdom), N.1.351 (South Africa), and P.1 (Brazil and Japan). To identify 15-mer peptides containing NP-specific minimal epitopes in GX-19N-vaccinated participants, we performed the IFN-γ ELISpot assay using a matrix of OLP spanning the entire NP or 15-mer individual peptides with PBMCs from four participants who mounted strong NP-specific T cell responses following GX-19N immunization. Subsequently, we compared the amino acids sequences of GX-19N and those of SARS-CoV-2 variants. These four participants induced T cell responses to multiple 15-mer peptides, indicating the induction of broad NP-specific T cell responses. (Supplementary Appendix 8) Importantly, we found that the amino acid sequences of 15-mer peptides containing T cell epitopes identified in GX-19N-vaccinated participants were identical (except for one 15-mer peptide) with those of SARS-CoV-2 variants (i.e., B.1.1.7, N.1.351, and P.1) (Figure 5D and Supplementary Appendix 8). These results imply that the NP-specific T cell responses induced by GX-19N can be cross-reactive with emerging SARS-CoV-2 variants.

## Discussion

We report the interim findings from a phase 1 clinical trial of two recombinant DNA vaccines administered to healthy adults. GX-19 and GX-19N were well tolerated and showed an acceptable safety profile without serious vaccine-related AEs. The overall incidence of solicited AEs in this study is lower than the frequencies observed in other platforms of licensed SARS-CoV-2 vaccines.^11–13^ The favorable safety profile in this study was comparable with the trial results of another DNA vaccine candidate for COVID-19.^14^

The GX-19N group showed stronger cellular responses than those of the GX-19 groups, which were similar to the T cell responses observed after natural COVID-19 infection. Recent studies have suggested the critical roles of SARS-CoV-2-specific T cells in the clearance of SARS-CoV-2 and protection from developing severe COVID-19. It has been demonstrated that among several immune parameters, the frequency of SARS-CoV-2-specific IFN-γ-producing CD8^+^ T cells showed the strongest inverse correlation with the severity of COVID-19.^15^ One study reported that SARS-CoV-2-specific polyfunctional T cells in the circulation of antibody negative convalescent patients presented asymptomatic and mild COVID-19^16^, suggesting in the absence of antibodies, a robust T cell response might provide immune protection against SARS-CoV-2.^17^ In another study, it has been shown that T cells contributed to protection of hosts with hematologic malignancy even when B cells were depleted by anti-CD20 therapy.^18^ Moreover, a study using a SARS-CoV-2 infection model of rhesus macaques showed that the depletion of CD8^+^ T cells in convalescent animals partially abolished the protective immunity against SARS-CoV-2 re-challenge.^19^ It was also reported that a T cell vaccination in the absence of neutralizing antibodies protected mice from SARS-CoV-2 infection.^20^

Furthermore, GX-19N not only induced S-specific T cell responses but also induced broad NP-specific T cell responses, which were also specific to SARS-CoV-2 variants. A key feature of GX-19N is that it contains a plasmid encoding NP, which is highly conserved (with >90% amino acid homology and fewer mutations over time) in diverse SARS-CoV-2 variants. To our knowledge, this is the first study reporting a phase 1 clinical trial of a SARS-CoV-2 DNA vaccine candidate that includes a non-spike protein-encoding DNA sequence. While escaping neutralizing antibody-based immunity is easily achieved via a few RBD mutations, escaping T cell-based immunity is far more complex because T cell epitopes are more abundant and widespread throughout the whole antigens.^21^ Therefore, considering the growing evidence suggesting that a T cell response is important for rapid virological control as well as long-term protection against SARS-CoV-2 infection, especially when neutralizing antibody titers decline, the fact that GX-19N induces broad T cell responses to highly conserved regions among SARS-CoV-2 variants indicates that this vaccine would be beneficial in inducing protective immunity against future SARS-CoV-2 variants.

Based on the binding and neutralizing antibody response assay results, GX-19N-induced antibody responses to SARS-CoV2 antigens are likely to be weaker than those induced by natural COVID-19 infection or commercialized mRNA-based or adenoviral vector-based vaccines^11–13^, even though immune responses are not directly compared in the same setting. Previous studies on DNA vaccines have determined that the number of doses required for effective vaccination is dependent on the antigen and virus types and how they interact with the immune system.^22^ One to two doses of the DNA vaccine are enough for eliciting sufficient neutralizing antibody responses to influenza virus, whereas three to four doses are needed to produce protective humoral responses against HIV.^23^ In a preclinical study of a plasmid DNA vaccine for SARS-CoV-2, it was reported that the magnitude of binding and neutralizing antibody responses was the highest after three doses.^22^ Further studies on humoral immunogenicity after another booster dose of GX-19N should be considered.

As this is a phase 1 interim analysis, we could not report persistent safety outcomes or vaccine-induced immune responses and the results do not permit a vaccine efficacy assessment. The interpretation of the results of this trial is also limited by the open-labeled, non-randomized design and small size. The participants in this trial were not ethnically diverse, and participants more susceptible to SARS-CoV-2, such as older individuals or individuals with comorbidities, were not included. However, this study has several strengths. This is the first trial to assess the safety and immunogenicity of a DNA vaccine that encodes both the S protein and the highly conserved NP. Given the concerns of decreased efficacy of the current COVID-19 vaccines against SARS-CoV-2 variants, we also performed an additional detailed assessment of T cell responses and analyzed the reactivity of vaccine-induced T cell responses to SARS-CoV-2 variants.

Our preliminary results indicate that the recombinant DNA vaccines for COVID-19, GX-19 and GX-19N are safe and that GX-19N is able to induce both humoral and cellular responses, more focused on T cell responses rather than neutralizing antibody responses. Importantly, GX-19N, which contains a plasmid encoding both the S protein and NP, showed a broad SARS-CoV-2-specific T cell response, which may allow cross-reactivity to emerging SARS-CoV-2 variants. Based on these safety and immunogenicity findings, GX-19N was selected as a vaccine candidate for phase 2 immunogenicity trials (NCT04715997).

## Supporting information

Supplementary Appendix will be used for the link to the file on the preprint site.

## Data Availability

The trials were registered with ClinicalTrials.gov (NCT044445389 and NCT04715997).

https://clinicaltrials.gov/ct2/show/NCT04715997

https://clinicaltrials.gov/ct2/show/NCT04445389

## Contributors

JYC, SHP and YCS are co-principal investigators of this trial, contributed to conceptualization of the trial and did research. JYA, JL, and YSS are co-first authors of this manuscript and contributed equally to the original draft writing. JWW contributed to conceptualization of the trial and study protocol. YGS, KHL, and JYC were responsible for the site work including participant recruitment, data collection, and follow up. MS, ECH, JWW verified the data. Safety data analysis and interpretation were performed by JYA, YJC and JYC. Immunogenicity tests were performed by JL, SHS, JWO, HYS, MK, JEK, JWY, and ECS. Immunogenicity data were analyzed by JYA, JL, YSS, JWO, JWW, ECS, SHP, and JYC. All authors contributed to manuscript reviewing and editing and approved the final version. All authors contributed to article preparation and decided to submit the article for publication.

## Data sharing

Deidentified individual participant data will be made available when the trial is complete, upon request directed to the corresponding author. Proposals should be sent to the corresponding authors, and data can be shared following proposal approval.

## Acknowledgements

This research was supported by the Korea Drug Development Fund funded by the Ministry of Science and ICT, Ministry of Trade, Industry, and Energy, and Ministry of Health and Welfare (HQ20C0016, Republic of Korea). The pathogen resources (NCCP43326) for this study were provided by the National Culture Collection for Pathogens. We thank the study participants, members of the trial management groups, site research staff and trial steering committee.

## References

1. Wibmer CK, Ayres F, Hermanus T, et al. SARS-CoV-2 501Y.V2 escapes neutralization by South African COVID-19 donor plasma. bioRxiv 2021.

2. Garcia-Beltran WF, Lam EC, Denis KS, et al. Circulating SARS-CoV-2 variants escape neutralization by vaccine-induced humoral immunity. medRxiv 2021.

3. Abdool Karim SS, de Oliveira T. New SARS-CoV-2 Variants - Clinical, Public Health, and Vaccine Implications. N Engl J Med 2021.

4. Eguia RT, Crawford KHD, Stevens-Ayers T, et al. A human coronavirus evolves antigenically to escape antibody immunity. PLoS Pathog 2021; 17(4): e1009453.

5. Agerer B, Koblischke M, Gudipati V, et al. SARS-CoV-2 mutations in MHC-I-restricted epitopes evade CD8(+) T cell responses. Sci Immunol 2021; 6(57).

6. Seo YB, Suh YS, Ryu JI, et al. Soluble Spike DNA Vaccine Provides Long-Term Protective Immunity against SARS-CoV-2 in Mice and Nonhuman Primates. Vaccines (Basel) 2021; 9(4).

7. Kim TJ, Jin HT, Hur SY, et al. Clearance of persistent HPV infection and cervical lesion by therapeutic DNA vaccine in CIN3 patients. Nat Commun 2014; 5: 5317.

8. Choi YJ, Hur SY, Kim TJ, et al. A Phase II, Prospective, Randomized, Multicenter, Open-Label Study of GX-188E, an HPV DNA Vaccine, in Patients with Cervical Intraepithelial Neoplasia 3. Clin Cancer Res 2020; 26(7): 1616–23.

9. KFaD A. Guideline for assessing severity of adverse events in clinical trials of vaccine. 2011.

10. UDoHaH S. Common terminology criteria for adverse events (CTCAE) version 5.0. 2017.

11. Mulligan MJ, Lyke KE, Kitchin N, et al. Phase I/II study of COVID-19 RNA vaccine BNT162b1 in adults. Nature 2020; 586(7830): 589–93.

12. Jackson LA, Anderson EJ, Rouphael NG, et al. An mRNA Vaccine against SARS-CoV-2 - Preliminary Report. N Engl J Med 2020; 383(20): 1920–31.

13. Folegatti PM, Ewer KJ, Aley PK, et al. Safety and immunogenicity of the ChAdOx1 nCoV-19 vaccine against SARS-CoV-2: a preliminary report of a phase 1/2, single-blind, randomised controlled trial. Lancet 2020; 396(10249): 467–78.

14. Andrade VM, Christensen-Quick A, Agnes J, et al. INO-4800 DNA Vaccine Induces Neutralizing Antibodies and T cell Activity Against Global SARS-CoV-2 Variants. bioRxiv 2021.

15. Rydyznski Moderbacher C, Ramirez SI, Dan JM, et al. Antigen-Specific Adaptive Immunity to SARS-CoV-2 in Acute COVID-19 and Associations with Age and Disease Severity. Cell 2020; 183(4): 996–1012 e19.

16. Sekine T, Perez-Potti A, Rivera-Ballesteros O, et al. Robust T Cell Immunity in Convalescent Individuals with Asymptomatic or Mild COVID-19. Cell 2020; 183(1): 158–68 e14.

17. de Candia P, Prattichizzo F, Garavelli S, Matarese G. T Cells: Warriors of SARS-CoV-2 Infection. Trends Immunol 2021; 42(1): 18–30.

18. Huang A, Bange E, Han N, et al. CD8 T cells compensate for impaired humoral immunity in COVID-19 patients with hematologic cancer. Res Sq 2021.

19. McMahan K, Yu J, Mercado NB, et al. Correlates of protection against SARS-CoV-2 in rhesus macaques. Nature 2020; 590(7847): 630–4.

20. Zhuang Z, Lai X, Sun J, et al. Mapping and role of T cell response in SARS-CoV-2-infected mice. J Exp Med 2021; 218(4).

21. Tarke A, Sidney J, Kidd CK, et al. Comprehensive analysis of T cell immunodominance and immunoprevalence of SARS-CoV-2 epitopes in COVID-19 cases. Cell Rep Med 2021; 2(2): 100204.

22. Almansour I, Macadato NC, Alshammari T. Immunogenicity of Multiple Doses of pDNA Vaccines against SARS-CoV-2. Pharmaceuticals (Basel) 2021; 14(1).

23. Wang S, Lu S. DNA immunization. Curr Protoc Microbiol 2013; 31: 18 3 1–3 24.

