## Supplementary Appendix will be used for the link to the file on the preprint site. for "Safety and immunogenicity of a recombinant DNA COVID-19 vaccine containing the coding regions of the spike and nucleocapsid proteins: Preliminary results from an open-label, phase 1 trial in healthy adults aged 19–55 years"

**Appendix 1. GX-19(N)-induced humoral and cellular responses in mice and monkeys**

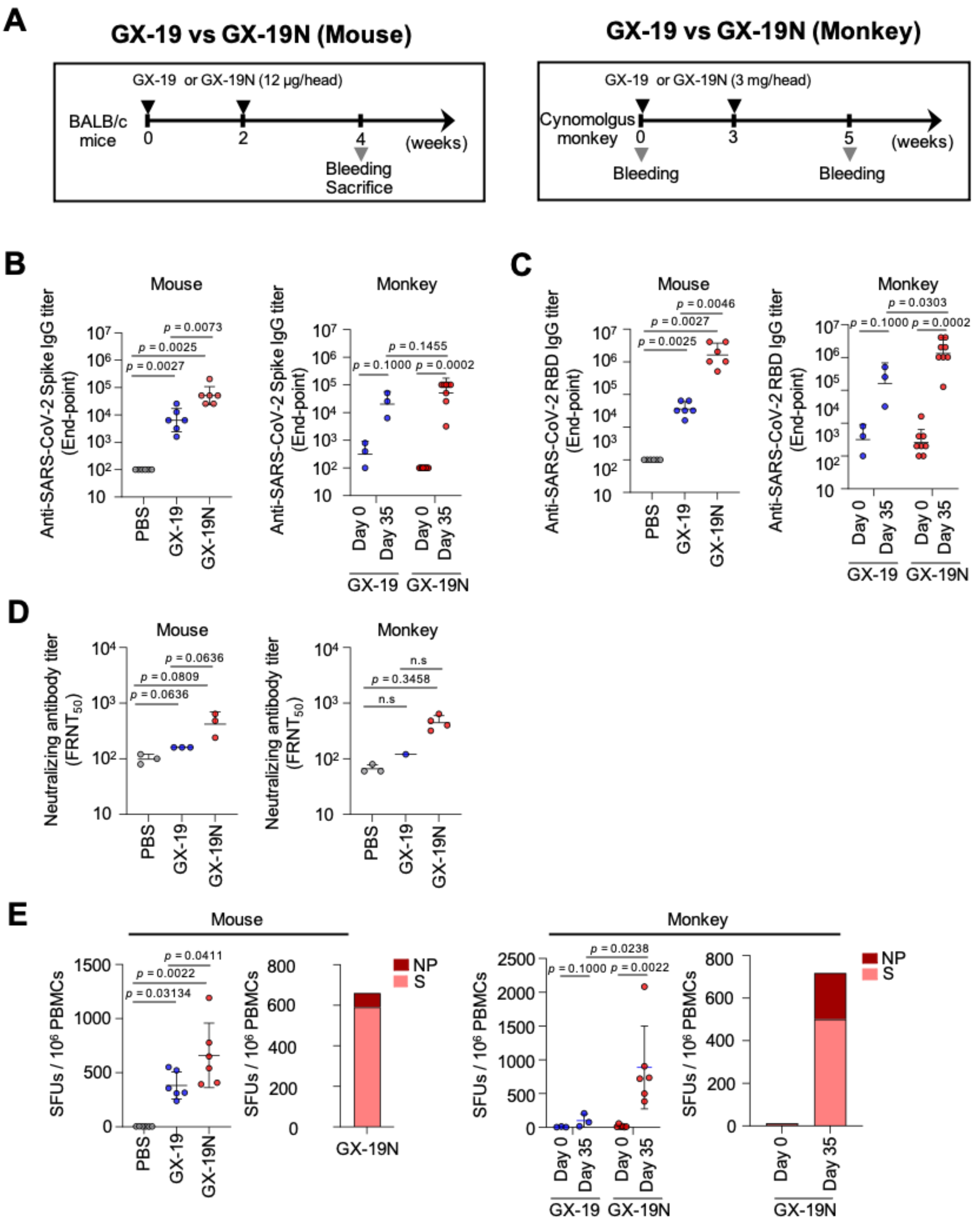

Timeline of pre-clinical trial (A), Results of anti-Spike IgG ELISA (B), Results of anti-SRBD IgG ELISA (C), Results of focus reduction neutralization test (D), Results of IFN-γ ELISPOT (E), S: spike, NP: nucleocapsid

**Appendix 2. Eligibility criteria of trial protocol**

Inclusion and Exclusion Criteria

1. Inclusion criteria

Candidates who meet all the criteria below will be determined to be eligible for inclusion in the study

1) Those who have been informed about the study and provide a voluntarily consent to participate in the study

2) Healthy adults (male or female) aged 19–49 years

3) Those with a body weight of 50–90 kg and body mass index (BMI) of 18.0–28.0 $kg/m^2$ at the time of screening

4) Those from whom blood and urine samples could be collected throughout the study period, including the EOS visit

2. Exclusion criteria

Candidates who meet any one of the criteria below will be determined to be ineligible and will be excluded from the study

1) Those with immune dysfunction, including immunodeficiency disorder, or family history of such conditions

2) Those with a history of a malignant disease within the past five years

3) Those with any surgery or dental treatment planned during the study period

4) Those who have received immunoglobulin or blood-derived products within three months prior to the vaccination or are scheduled to receive them during the study period

5) Those who have been dependent on antipsychotic drugs and narcotic analgesics within six months prior to the vaccination

6) Those who test positive on serological test during the screening period [hepatitis B test, hepatitis A test, human immunodeficiency virus test; hepatitis C test; and SARS-CoV-2 (COVID-19 IgG Ab) test]

7) Those who have a history or are suspected of drug abuse in the last 12 months prior to the vaccination

8) Those who continue to drink alcohol (exceeding 21 units/week) or have a history of alcohol abuse

9) Those with a history of allergy or SAEs to any antibiotics, nonsteroidal anti-inflammatory drugs (NSAIDs), or drug or vaccine containing ingredients of GX-19 or same class of ingredients

10) Those with a history of hypersensitivity to vaccination, such as those with Guillain–Barre syndrome

11) Those with current or previous history of other biliary, renal, nervous (central or peripheral), respiratory (asthma, pneumonia, etc.), endocrine (uncontrolled diabetes, hyperlipidemia, etc.), cardiovascular (congestive heart failure, coronary artery disease, myocardial infarction, uncontrolled hypertension, etc.), hemato-oncologic, urinary, psychological, musculoskeletal, or immune (rheumatic arthritis, systemic lupus erythematosus, immunodeficiency disorder, etc.) disorders, which the principal investigator determines to be clinically significant to make participation in the study impossible

12) Hemophiliacs or people using anticoagulants who are at a risk of serious bleeding from IM injection

13) Those who have been in close contact with a COVID-19 patient or identified as a confirmed or suspected COVID-19 patient or symptomatic patient under investigation prior to vaccination or those with a previous history of confirmed SARS or MERS infection

14) Those with acute fever with temperature above 37·5°C, coughing, breathing difficulty, chills, muscle ache, headache, sore throat, loss of smell, or loss of taste within 72 hours prior to the vaccination

15) Those who have a history of other vaccination(s) within 28 days prior to the vaccination or are scheduled to receive other vaccinations during the study period

16) Those who have taken an immunosuppressant or immune-modifying drug within three months prior to the vaccination

① Including but not limited to azathioprine, cyclosporin, interferon, G-CSF, tacrolimus, everolimus, sirolimus, cyclophosphamide, 6-mercaptopurine, methotrexate, rapamycin, and leflunomide

② Those who have taken systemic steroids for 14 days or more within three months prior to screening (for prednisolone, continuously taking a maximum dose of ≥10 mg/day for more than 14 days would be considered high dose and participation in this study would be restricted)

17) Those who have received other investigational drug(s) while participating in another clinical trial or bioequivalence study within six months prior to vaccination

18) Women who are pregnant or breastfeeding; however, participation is possible if breastfeeding is discontinued prior to participation in the study (women of childbearing age† must test negative in serum pregnancy test during the screening period prior to the start of the study)

†Women of childbearing age: Women who have started menarche and have not yet reached menopause (at least 12 months elapsed after non-therapy-induced amenorrhea) and have not received any surgical sterilization procedure (oophorectomy and/or hysterectomy)

19) Women of childbearing age who did not consent to the use of effective contraception† (condoms, contraceptive diaphragms, and/or intrauterine contraceptive devices after completing the consent form) during the study period

†Refer to section 9.2.1 for effective contraception

20) Those deemed ineligible by the investigator based on other clinically significant medical or psychiatric findings

2.1. Exclusion criteria: Definition of effective contraception

Women of childbearing age who wish to participate in the study but do not consent to use effective contraception during the study period may not be eligible to participate in the study. Women who have not received any surgical sterilization procedure or are not in menopause (at least 12 months elapsed after non-therapy-induced amenorrhea) must avoid becoming pregnant. Women who have an active sex life with a male partner and could become pregnant must practice abstinence (refraining from intercourse with the opposite sex) or use contraception. While refraining from sexual relationship could be recognized as a form of contraception, temporary abstinence and coitus interruptus are not permissible contraceptive methods. The effective contraceptive methods defined in this study are as follows, and they could be divided broadly into barrier and non-barrier methods. For barrier methods, dual protection must be used.

- Non-barrier methods:

① Intrauterine hormonal device and intrauterine device (IUD): Copper IUD, hormone-releasing IUD, copper T IUD, levonorgestrel-releasing IUD (e.g., Mirena®), ethinyl estradiol/etonogestrel intravaginal ring, etc.

② Oral contraceptives: Regular and low-dose complex oral contraceptives, norelgestromin/ethinyl estradiol transdermal system, Cerazette, etc.

③ Contraceptive injections: Etonogestrel implant, medroxyprogesterone injection, hormone-based methods (e.g., Depo-Provera), etc.

④ Others: Spermicidal foam, sponge, film, hormone patch, etc.

- Barrier methods: Diaphragm, cervical cap, cervical shield, male condom, female condom, etc.

Female subjects of childbearing age and male subjects with a female sex partner of childbearing age must opt for contraception using any of the methods described above from the day of a priming vaccination to 90 days after the boosting vaccination. All subjects must also refrain from egg/sperm donation. The investigator must be notified immediately if a female subject or the female partner of a male subject becomes pregnant during the study period or after the completion of the study (at least 90 days). If a subject becomes pregnant during the study period, she will receive no additional vaccination and will be dismissed from the study.

When a female subject reports her own pregnancy or a male subject reports the pregnancy of his female partner, the investigator and sub-investigators may collect data regarding the pregnancy and/or childbirth and plan a schedule for follow-up visits when necessary.

**Appendix 3. Study procedures and assessments**

| Schedule | Screening | Priming vaccination | Telephone† Visit | Boosting vaccination | Blood Sampling Visit | Blood Sampling Visit | Blood Sampling Visit | End-of-study Visit |
|---|---|---|---|---|---|---|---|---|
| Day | -28 ~ 0 | 1 | 8+3D | 29±3D | 43+3D | 57+3D | 24W±14D | 52W±14D |
| Visit / Item | V1 | V2 | V3 | V4 | V5 | V6 | V7 | V8[12)] |
| Obtain informed consent | ● | | | | | | | |
| Demographic investigation | ● | | | | | | | |
| Disease/surgery history taking[1)] | ● | | | | | | | |
| Inclusion/exclusion criteria | ● | ● | | | | | | |
| Physical examination[2)] | ● | ● | ●† | ● | ● | ● | ● | ● |
| Vital signs and body weight[2)] | ● | ● | ●† | ● | ● | ● | ● | ● |
| Chest X-ray examination[3)] | ● | | | | | | | ● |
| ECG[4)] | ● | ● | | ● | | | | ● |
| Randomization[5)] | | ● | | | | | | |
| **Blood Sampling Items and Schedule** | | | | | | | | |
| Hematology and blood chemistry test[6)] | ● | ● | ●† | ● | ● | ● | ● | ● |
| Serology test[7)] | ● | | | | | | | |
| Coagulation test[8)] | ● | ● | | ● | | | | |
| Urinalysis[9)] | ● | ● | ●† | ● | ● | ● | ● | ● |
| Pregnancy test[10)] | ● | | | | | | | ● |
| Immunogenicity blood sampling | ● | ● | | | ● | ● | ● | ● |
| **Related to IP Administration** | | | | | | | | |
| Vaccination and pain assessment[11)] | | ● | | ● | | | | |
| Previous and concomitant medication[12)] | ● | ● | ● | ● | ● | ● | ● | ● |
| Distribution of subject diary[13)] | | ● | | ● | | | | |
| Retrieval of subject diary and assessment[14)] | | | | ● | | ● | | |
| AE monitoring[15)] | | ● | ● | ● | ● | ● | ● | ● |
| Telephone monitoring[16)] | | | ● | | | | | |

† All of the subjects in Phase 1 will be checked daily over the phone for AEs for up to seven days from the day of the priming vaccination, and safety-related data will be collected. First three subjects enrolled in each experimental group in GX-19 trial and all of the subjects in GX-19N trial will undergo safety assessment (tests if necessary) through an outpatient visit, at 8 days (+3D) after the priming vaccination.

‡ If V1 and V2 take place on the same day, physical examination, vital signs and body weight measurement, electrocardiography (ECG) before the vaccination, hematology and blood chemistry test, coagulation test, and urinalysis results from V1 may be used as results for V2.

1) Previous and current medical history of each subject is obtained by inquiry and from past medical records within one year of the screening visit.

2) Physical examination and measurement of vital signs and body weight are performed prior to the vaccination.

3) Examination may be performed again if deemed necessary by the investigator.

4) ECG is performed during the screening (V1), end-of-study (V8) visits, and within 35 min before and after the vaccination only for the Subjects enrolled in Phase 1.

5) Randomization and vaccination (including placebo) apply only to phase 2a.

6) Hematology test: WBC differential count (neutrophil, lymphocyte, monocyte, eosinophil, and basophil), RBC, Hb, Hct, and platelet count (including absolute neutrophil counts and absolute lymphocyte counts). If the results of a specific test deviate from the normal level, the investigator may ask the subject for a re-visit to repeat only that test. Blood chemistry test: Total protein, albumin, creatinine, BUN or urea, total bilirubin, alkaline phosphatase, ALT, AST, GGT, glucose, calcium, magnesium, phosphorus, chloride, sodium, potassium, bicarbonate, LDH, amylase, lipase, and CRP. If the results of a specific test deviate from the normal level, the investigator may ask the subject for a re-visit to repeat only that test.

7) HBV (HBsAg), HAV (anti-HAV IgM), HIV antigen/antibody combination assay, HCV (anti-HCV), and SAS-CoV-2 Ab are performed

8) Coagulation test: Includes aPTT, PT, and INR. The test is performed during the visits for the vaccination. If screening (V1) and the priming vaccination (V2) visit are less than two weeks apart, the test results from V1 could be used for V2.

9) Urinalysis: RBC, WBC, pH, blood, glucose, urobilinogen, ketone, albumin, and bacteria

10) Pregnancy test by blood will be performed only in women in the childbearing age. Women who have received any surgical sterilization procedure (oophorectomy and/or hysterectomy) or menopausal women (at least 12 months elapsed after nontherapy-induced amenorrhea) are exempted.

11) Immunogenicity blood Sampling is performed to collect PBMC and Serum to evaluate the antibody response during All visits except V3 and V4. However, it can be optionally performed during Screening visit (V1) according to the individual visit schedules. Blood sampling must be performed before the vaccination during V2 and V4.

12) Pain assessment by using VAS is performed immediately after the vaccination and after 10 minutes of the vaccination.

13) Previous medication used within 12 weeks from the screening visit and concomitant medication to be used during the study period must be listed. Concomitant medications other than those prescribed by the principal investigator to treat AEs are not permitted.

14) The subject diary will be distributed at the time of the priming and the boosting vaccination. Each subject will personally list any AEs experienced over approximately four weeks. The diary will be retrieved during V4 and V6 and assessed. Subsequently, telephone or unscheduled visits will be permitted at any time, during which data regarding any AEs that may have occurred could be collected.

15) Information about AEs will be collected during scheduled visits after the vaccination and telephone visit.

16) Within 7–10 days after the Priming vaccination, a trained person designated by the principal investigator will call the subject to assess safety and notify the subject of the schedule for the following visit.

17) The end-of-study visit (V8) will be at 52 weeks after the priming vaccination. If necessary, follow-up visits may be required for long-term safety and immunogenicity assessment. Moreover, subjects who dropout from the study must go through a procedure equivalent to the end-of-study visit

### Appendix 4. Methods of determination of immunogenicity

SARS-CoV-2 Spike protein Ab ELISA

Ninety-six-well ELISA plates (Nunc Immunomodule, Thermo Fisher Scientific) were coated room temperature overnight with 200 ng per well of the purified recombinant SARS-CoV-2 S protein (Acro biosystem) in coating buffer (Carbonate-Bicarbonate buffer pH9.6, Sigma Aldrich). The plates were then blocked with 200 μL of 2% skim milk in PBS buffer and incubated at s 37℃ incubator for 2 h. Each serum sample was serially diluted 1:2 in sample dilution buffer (PBS-1% BSA-0.05% Tween 20-10% Skim milk) starting at a dilution of 1:10. 100 μL of each dilution was added and incubated for 12~16 hrs at s 37℃ incubator. 100 μL of Goat-Anti-Human IgG Fc Fragment HRP, diluted 1 to 10,000 in PBS-1% BSA buffer, were added and incubated for 1 h at a 37℃ incubator. Development was performed using TMB substrate (Surmodics). Reactions were stopped with 1N sulfuric acid and absorbance was measured at 450 nm. Between each step, samples were washed 5 times with 300 μL of PBS-0·05 Tween including prior to development. The titer was obtained by calculating each plate cutoff level (OD of plate negative control x 0·178).

SARS-CoV-2 RBD Ab ELISA

The binding antibody responses against the receptor binding domain (RBD) with ELISA kit manufactured by Bionote. A dilution of 1:1 was the positivity cut off value for ELISA. Samples were serially diluted 1:2 in sample dilution buffer (PBS-1% BSA) starting at a dilution of 1:1. 50 μL of sample dilution buffer was added each well and 50 μL of each dilution was added and incubated for 2 h at a 37℃ incubator. 100 μL of Enzyme conjugate, diluted to 0.1 μg/mL in enzyme conjugate diluent, were added and incubated for 1 h at a 37℃ incubator. Development was performed using TMB substrate and reactions were stopped with 1N sulfuric acid and absorbance was measured at 450 nm. Between each step, samples were washed 5 times with 300 μL of wash buffer including prior to development. The titer was obtained by calculating the plate cutoff level (0·062).

Pseudovirus neutralization assay

Neutralization efficiency of anti-SARS-CoV-2 spike antibody in sera from vaccinees or convalescent patients was evaluated using SARS-CoV-2 Spike (S)-pseudotyped murine leukemia virus (MLV) retrovirus (SARS2-Spp). The packaging plasmid pUMVC (Addgene plasmid #8449) (2) and the transfer plasmid pBABE-puro-NanoLuc, which was constructed by inserting the NanoLuc-coding gene into pBABE-puro (Addgene plasmid #1764), were used to prepare the SARS2-Spp by transfection of these plasmids into a HEK293T cell line (HEK293T/Spike) stably expressing a C-terminal 19 amino acid deleted SARS-CoV-2 spike protein (Wuhan-1 strain), as described previously. SARS2pp titer was determined by real-time RT-PCR using the Takara Retrovirus Titer Set (Takara Bio, Kusatsu, Shiga, Japan) and aliquots were frozen at −80°C until used for transduction experiments.

Equal volumes (400 μL each) of diluted sera (starting with 1 to 2 with four twofold dilutions for pre-vaccination sera, 1 to 4 with ten twofold dilutions for post-vaccination sera, or 1 to 40 with eight twofold dilutions for convalescent sera from patients) and diluted SARS2pp, which yields ~$10^6$ nanoluciferase reads (relative light units, RLU), were mixed (n = 3) and incubated for 1 h at 37°C. Then HEK293T cells stably expressing hACE2 (HEK293T/hACE2), which were grown for 48 h following seeding at 1 x $10^5$/well on 12-well plates, were transduced with the pseudotyped virus. Following media change at 12 h post-transduction, cells were further cultivated for 36 h and lysed in 115 μL of Glo Lysis Buffer (Promega, Madison, WI, USA). The resulting cell lysates (75 μL) were transferred to white, flat bottom 96-well microplates (Greiner Bio-One, Kremsmünster, Austria) for luciferase assay using the Nano-Glo Luciferase Assay system (Promega). The 50% inhibitory dilution (ID50), which was defined as the serum dilution causing a 50% reduction in luciferase activity compared to the wells displaying no inhibition, was determined by plotting a dose-response curve (log10 nanoluciferase activity vs. dilution) and four-parameter non-linear regression analysis using GraphPad prism 8·0 (GraphPad Software Inc., San Diego, CA, USA).

Live virus neutralization assay.

Vero cells were seeded on 96-well plate (NUNC cat#167008) and incubate for 16 h at 37°C 5% $CO_2$. Sera from vaccinee were heat-inactivated by incubation at 56°C for 30 min. The sera were serially diluted, mixed with SARS-CoV-2 (450 plaque-forming unit) and the mixture was incubated for 30 min at 37°C 5% $CO_2$. After washing the Vero cells using serum-free DMEM (Invitrogen, cat# 11995065), the virus-sera mixture was treated into the Vero cells and the mixture treated Vero cells were incubated for 5 h at 37°C and 5% $CO_2$. After removal of the treated mixture supernatant, the cells were washed using 100µl of phosphate buffered saline (PBS) (Gibco, cat# 10010-023) and fixed using 300 µL of 10% formalin solution (Sigma, cat# F8775) by incubation for 16 h at 4°C. After washing, the Vero cells were permeabilized by adding 100 µL of ice-cold 100% methanol (Sigma, cat# D7), incubated for 10 min at room-temperature, and washed using 100µl PBS. The cells were blocked by incubation with 100 µL of blocking buffer [0·5% normal goat serum (Abcam, cat# Ab7481) supplemented with 0·1% Tween 20 (GenDEPOT, cat# T9100-100) and 1% (w/v) Bovine serum albumin (Sigma, cat#A3803-100G) in PBS)] for 30 min at room temperature. Then the blocked cells were incubated with 3,000-fold diluted 100 µL of anti-SARS-CoV-2 NP rabbit mAb (Sino Biological, cat# 40143-R001) for 1h at 37°C. After washing the cells using 200 µL of PBS containing 0·1% Tween 20, the Vero cells were incubated with 2,000-fold diluted goat anti-rabbit IgG-HRP (Bio-Rad, cat# 170-6515) solution for 1h at 37°C. After washing, the Vero cells were incubated with 30 µL of TrueBlue solution for 30 min at room temperature. After removal of the TrueBlue solution, the cells were air dried completely. The numbers of focus of each well were read using Cytation7 (Cellular Biotek) and the neutralizing Ab titers were calculated using Microsoft Excel and SoftMax (Version 5.4.1.).

ELISpot assay

The human IFN-γ ELISpot assay (Mabtech) was performed as recommended by the manufacturer. PBMCs were isolated from whole blood by Ficoll-Hypaque density gradient centrifugation and resuspended ($0.3 \times 10^6$ cells per well) in RPMI medium. The PBMCs were stimulated with SARS-CoV-2 S or NP OLP pools for 24 h at 37° C in a 5% $CO_2$ atmosphere. Spots were enumerated using an automated ELISpot reader (AID GmbH,) and the number of specific spots was calculated by subtracting the number of spots in negative control wells from the number of spots in OLP pool-stimulated wells.

To selectively evaluate vaccine-induced SARS-CoV-2-specific T cell responses, the T-SPOT (Oxford Immunotec) was performed as recommended by the manufacturer. In this T-SPOT assay, PBMCs from participants were stimulated with OLPs spanning S and NP that specific sequences with high homology to endemic (non SARS-CoV-2) coronaviruses removed were used as stimulators for 24 h at 37° C in a 5% $CO_2$ atmosphere.

Overlapping peptides

OLPs were synthesized as 15-mers that overlapped by 11 amino acids to cover the whole amino acid sequence of SARS-CoV-2 S and NP proteins encoded by GX19 and GX-19N. Lyophilized peptides were solubilized in 5% dimethyl sulfoxide (DMSO; Sigma Aldrich). The concentration of each peptide in the pools was 25 µg $mL^{-1}$, which was finally diluted to 1 µg $mL^{-1}$ for stimulation of peripheral blood mononuclear cells (PBMCs) to examine vaccine-induced T cell responses in participants.

**Appendix 5. Incidence of treatment-emergent adverse events (TEAEs) by system organ class (SOC) and preferred term (PT)**

| TEAEs by SOC and PT | GX-19 1·5mg (N=20) | GX-19 3·0mg (N=20) | GX-19N 3·0mg (N=21) |
|---|---|---|---|
| **Subjects with TEAEs** | 12(60·00) [33] | 8(40·00) [16] | 12(57·14) [31] |
| **General disorders and administration site conditions** | 11(55·00) [21] | 4(20·00) [5] | 6 (28·57) [16] |
| Chills | 0 | 0 | 1 (4·76) [1] |
| Fatigue | 3 (15·00) [4] | 0 | 0 |
| Injection site erythema | 2 (10·00) [2] | 0 | 1 (4·76) [1] |
| Injection site oedema | 1 (5·00) [1] | 0 | 0 |
| Injection site pain | 6 (30·00) [8] | 3 (15·00) [3] | 6 (28·57) [14] |
| Injection site pruritus | 5 (25·00) [6] | 2 (10·00) [2] | 0 |
| **Nervous system disorders** | 4(20·00) [6] | 0 | 2 (9·52) [2] |
| Cubital tunnel syndrome | 0 | 0 | 1 (4·76) [1] |
| Headache | 3(15v00) [5] | 0 | 1 (4·76) [1] |
| Paresthesia | 1(5·00) [1] | 0 | 0 |
| **Musculoskeletal and connective tissue disorders** | 1(5·00) [2] | 1(5·00) [2] | 2 (9·52) [2] |
| Back pain | 0 | 0 | 1 (4·76) [1] |
| Myalgia | 1(5·00) [2] | 0 | 1 (4·76) [1] |
| Rotator cuff syndrome | 0 | 1(5·00) [1] | 0 |
| Somatic dysfunction | 0 | 1(5·00) [1] | 0 |
| **Gastrointestinal disorders** | 2 (10·00) [2] | 1(5·00) [1] | 0 |
| Dyspepsia | 1(5·00) [1] | 0 | 0 |
| Hematochezia | 1(5·00) [1] | 0 | 0 |
| Stomatitis | 0 | 1(5·00) [1] | 0 |
| **Skin and subcutaneous tissue disorders** | 0 | 1 (5·00) [1] | 3 (19·05) [4] |
| Dermatitis | 0 | 1 (5·00) [1] | 0 |
| Pruritus | 0 | 0 | 1 (4·76) [1] |
| Rash | 0 | 0 | 2 (9·52) [2] |
| Urticaria | 0 | 0 | 1 (4·76) [1] |
| **Infections and infestations** | 0 | 2 (10·00) [2] | 1 (4·76) [2] |
| Appendicitis | 0 | 1 (5·00) [1] | 0 |
| Folliculitis | 0 | 0 | 1 (4·76) [2] |
| Vaginal infection | 0 | 1 (5·00) [1] | 0 |
| **Investigations** | 0 | 0 | 1 (4·76) [2] |
| Alanine aminotransferase increased | 0 | 0 | 1 (4·76) [1] |
| Aspartate aminotransferase increased | 0 | 0 | 1 (4·76) [1] |
| **Reproductive system and breast disorders** | 0 | 2 (10·00) [2] | 1 (4·76) [2] |
| Dysmenorrhoea | 0 | 1 (5·00) [1] | 1 (4·76) [2] |
| Premenstrual syndrome | 0 | 1 (5·00) [1] | 0 |
| **Hepatobiliary disorders** | 0 | 0 | 1 (4·76) [1] |
| Cholecystitis acute | 0 | 0 | 1 (4·76) [1] |
| **Psychiatric disorders** | 1 (5·00) [1] | 0 | 0 |
| Depression | 1 (5·00) [1] | 0 | 0 |
| **Eye disorders** | 1 (5·00) [1] | 1 (5·00) [2] | 0 |
| Blepharospasm | 0 | 1 (5·00) [2] | 0 |
| Visual impairment | 1 (5·00) [1] | 0 | |

MedDRA version: 24.0.
Denominator of percentage is the number of subjects in each cohort.
Adverse events are displayed as number of subjects (percentage of subjects) [number of events]

**Appendix 6. Incidence of adverse drug reaction (ADR) by system organ class (SOC) and preferred term (PT)**

| Subjects with ADRs | GX-19 1·5mg (N=20) | GX-19 3·0mg (N=20) | GX-19N 3·0mg (N=21) |
|---|---|---|---|
| **ADRs by Severity** | 11 (55·00) [24] | 4 (20·00) [5] | 10 (47·62) [22] |
| Mild | 11 (55·00) [23] | 4 (20·00) [5] | 10 (47·62) [22] |
| Moderate | 1 (5·0) [1] | 0 | 0 |
| Severe | 0 | 0 | 0 |
| **ADRs by SOC and PT** | | | |
| **General disorders and administration site conditions** | 11 (55·00) [21] | 4 (20·00) [5] | 6 (28·57) [16] |
| Chills | 0 | 0 | 1 (4·76) [1] |
| Fatigue | 3 (15·00) [4] | 0 | 0 |
| Injection site erythema | 2 (10·00) [2] | 0 | 1 (4·76) [1] |
| Injection site oedema | 1 (5·00) [1] | 0 | 0 |
| Injection site pain | 6 (30·00) [8] | 3 (15·00) [3] | 6 (28·57) [14] |
| Injection site pruritus | 5 (25·00) [6] | 2 (10·00) [2] | 0 |
| **Skin and subcutaneous tissue disorders** | 0 | 0 | 2 (9·52) [2] |
| Rash | 0 | 0 | 2 (9·52) [2] |
| **Musculoskeletal and connective tissue disorders** | 1 (5·00) [2] | 0 | 1 (4·76) [1] |
| Myalgia | 1 (5·00) [2] | 0 | 1 (4·76) [1] |
| **Nervous system disorders** | 1 (5·00) [1] | 0 | 1 (4·76) [1] |
| Headache | 0 | 0 | 1 (4·76) [1] |
| Paresthesia | 1 (5·00) [1] | 0 | 0 |
| **Investigations** | 0 | 0 | 1 (4·76) [2] |
| Alanine aminotransferase increased | 0 | 0 | 1 (4·76) [1] |
| Aspartate aminotransferase increased | 0 | 0 | 1 (4·76) [1] |

MedDRA version: 23.0.
Denominator of percentage is the number of subjects in each cohort.
Adverse events are displayed as number of subjects (percentage of subjects) [number of events]

**Appendix 7. Results of ELISpot and T-SPOT in GX-19N-vaccinated participants**

| | ELISPOT (SFUs / $10^6$ PBMCs) | | | T-SPOT (SFUs / $10^6$ PBMCs) | | |
|---|---|---|---|---|---|---|
| **Vaccinee** | **Pre** | **Week 6** | **Ratio** | **Pre** | **Week 6** | **Ratio** |
| **19Na** | 698·3 | 363·3 | 0·5 | 28·0 | 64.0 | 2·3 |
| **19Nb** | 290·0 | 1550·0 | 5·3 | | N/D | |
| **19Nc** | 270·0 | 3663·3 | 13·67 | | N/D | |
| **19Nd** | 835·0 | 546·67 | 0·7 | 272·0 | 248.0 | 0·9 |
| **19Ne** | 176·7 | 2756·7 | 15·6 | | N/D | |
| **19Nf** | 863·3 | 791·7 | 0·9 | 120·0 | 580.0 | 4·8 |
| **19Ng** | 605·0 | 2006·7 | 3·3 | | N/D | |
| **19Nh** | 1016·7 | 3250·0 | 3·2 | | N/D | |
| **19Ni** | 420·0 | 2463·3 | 5·9 | | N/D | |
| **19Nj** | 1053·3 | 3420·0 | 3·3 | | N/D | |
| **19Nk** | 63·3 | 1040·0 | 16·4 | | N/D | |
| **19Nl** | 330·0 | 985·0 | 3·0 | | N/D | |
| **19Nm** | 113·3 | 1351·7 | 11·9 | | N/D | |
| **19Nn** | 496·7 | 1271·7 | 2·6 | | N/D | |
| **19No** | 415·0 | 2520·0 | 6·1 | | N/D | |
| **19Np** | 523·3 | 695·0 | 1·3 | | N/D | |
| **19Nq** | 113·3 | 135·0 | 1·3 | 28·0 | 196·0 | 7·0 |
| **19Nr** | 61·7 | 516·7 | 8·4 | | N/D | |
| **19Ns** | 326·7 | 756·7 | 2·3 | | N/D | |
| **19Nt** | 358·3 | 293·3 | 0·8 | | N/D | |

N/D, not determined; SFU, spot-forming units; PBMCs, peripheral blood mononuclear cells.

**Appendix 8. Comparison of amino acids sequence of 15-mer peptides containing NP-specific epitopes for GX-19N-vaccinated participants with those of SARS-CoV-2 variants**

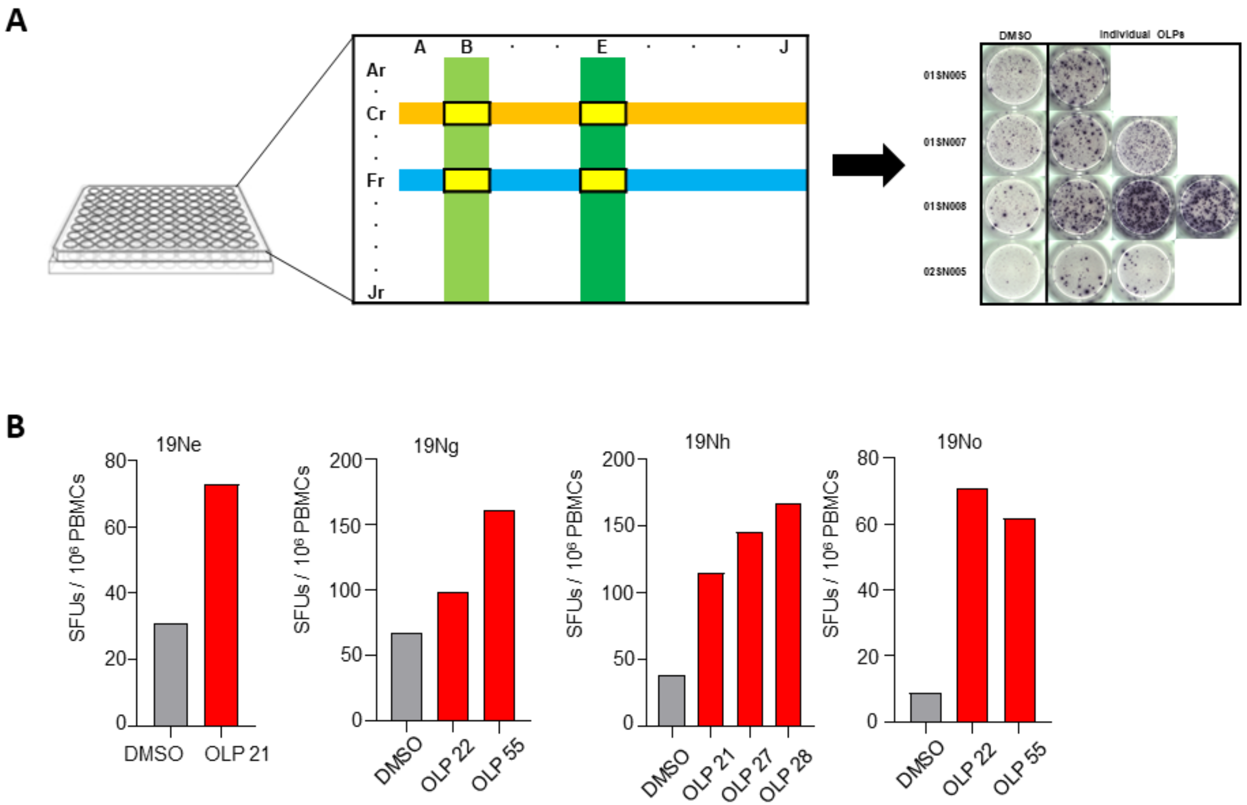

Strategy to determine 15-mer peptides containing NP-specific epitopes for GX-19-vaccinated participants (A), Results of IFN-γ ELISPOT assay using 15-mer individual peptide (B), NP, nucleocapsid protein
